# Genome-wide Association Study Meta-analysis of Neurofilament light (NfL) levels in blood reveals novel loci related to neurodegeneration

**DOI:** 10.1101/2022.12.14.22283446

**Authors:** Shahzad Ahmad, Mohammad Aslam Imtiaz, Aniket Mishra, Ruiqi Wang, Marisol Herrera-Rivero, Joshua C Bis, Myriam Fornage, Gennady Roshchupkin, Edith Hofer, Mark Logue, WT Longstreth, Rui Xia, Vincent Bouteloup, Thomas Mosley, Lenore Launer, Michael Khalil, Jens Kuhle, Robert A. Rissman, Genevieve Chene, Carole Dufouil, Luc Djoussé, Michael J. Lyons, Kenneth J. Mukamal, William S. Kremen, Carol E. Franz, Reinhold Schmidt, Stephanie Debette, Monique M.B. Breteler, Klaus Berger, Qiong Yang, Sudha Seshadri, N. Ahmad Aziz, Mohsen Ghanbari, M. Arfan Ikram

## Abstract

**Background:** Neurofilament light chain (NfL) levels in circulation have been established as a sensitive biomarker of neuro-axonal damage across a range of neurodegenerative disorders. Elucidation of the genetic architecture of blood NfL levels and its genetic correlation with neurological traits could therefore provide new insights into shared molecular mechanisms underlying neurodegenerative disorders.

**Methods:** To identify the genetic variations underlying blood NfL levels, we conducted an ancestry-specific meta-analyses of genome-wide association studies (GWAS) based on 18,532 participants from 11 cohorts of European and 1142 participants (3 cohorts) of African-American ancestry. In the post-GWAS analyses, we performed expression quantitative trait loci (eQTL) analysis, LD-regression, and genetic risk score (GRS) association analysis with neurological traits.

**Results:** In the European ancestry GWAS meta-analysis, we identified two genome-wide significant (*P* < 5x10^−8^) loci at 16p12 (*UMOD)*, and 17q24 (*SLC39A11*). In the African-American ancestry GWAS meta-analysis, we identified three novel loci at 1q43 (*FMN2*), 12q14, and 12q21. Genetic correlation based on the European ancestry meta-analysis with neurological traits showed a strong genetic correlation of NfL with Alzheimer’s disease(AD) (*r*_*g*_ = 0.32, *P* = 1.74x10^−6^), total-tau (*r*_*g*_ = 2.01, *P* = 1.03x10^−6^), amyloid-beta (Aβ)-40 (*r*_*g*_ = 0.80, *P* = 6.92x10^−6^), and Aβ-42 (*r*_*g*_ = 1.03, *P* = 4.39x10^−5^). A higher genetic risk score based on NfL-associated genetic variants was also related to increased plasma levels of total-tau (*P* = 1.97x10^−4^), Aβ-40 (*P* = 2.24x10^−5^), Aβ-42 (*P* = 2.92x10^−4^) in the Rotterdam Study.

**Conclusion:** This large-scale GWAS meta-analysis revealed multiple novel genetic loci of NFL levels in blood in participants from European and African-American ancestry. Significant genetic correlation of genes underlying NfL with AD, Aβ-42, and total-tau may indicate a common underlying pathway of neurodegeneration.

## Introduction

Blood levels of the neurofilament light chain (NfL) have emerged as a robust biomarker of neuro-axonal injury and are increased in a range of neurodegenerative disorders including Alzheimer’s disease (AD), Parkinson’s disease (PD), amyotrophic lateral sclerosis (ALS), and multiple sclerosis(1). NfL proteins are expressed in the cytoplasm of neurons where they confer structural stability to the cytoskeleton of neurons(1, 2). Under normal physiological conditions, NfL proteins are continuously released from the axoplasm into circulation in an age-dependent manner(3), whereas neuro-axonal damage has been associated with increased release of NfL in the neuronal extracellular space(3, 4). NfL proteins may diffuse into the cerebrospinal fluid (CSF) and circulation, thereby acting as biomarkers of neuro-axonal injury and neurodegeneration(5, 6). Identifying the genetic basis of the NfL in blood could therefore provide a better understanding of the biological pathways underlying axonal damage and facilitate identification of shared molecular mechanisms contributing to neuronal loss across neurodegenerative disorders.

Previously, two modest size genome-wide association studies (GWAS) were performed to identify the genetic variants associated with plasma, and CSF levels of NfL in the Alzheimer’s Disease Neuroimaging Initiative (ADNI) cohort(7, 8). The GWAS on the CSF levels of NFL suggested two genome-wide significant association of variants near the *ADAMTS1* gene at chromosome 21(7), and sub-threshold associations of *LUXP2* and *GABRB2* genes with plasma levels of NFL(8). To uncover the underlying genetic factors of blood NfL levels, studies with substantially larger sample sizes across different ethnicities are warranted.

In the current study, we therefore performed a GWAS meta-analysis based on the findings from 11 cohorts of the Cohorts for Heart and Aging Research in Genomic Epidemiology (CHARGE) consortium including people from both European and African-American ancestry. Furthermore, we performed a range of post-GWAS investigations, including expression quantitative trait loci (eQTLs) lookups, pathway enrichment analysis, and linkage disequilibrium (LD) regression analysis. Based on the identified genetic variants of NfL, we calculated a genetic risk score (GRS) and assessed its association with the incidence of AD, and other AD-related endo-phenotypes using individual levels data from the Rotterdam Study cohort.

## Methods

### Study populations

The current study includes 18532 participants of European and 1142 participants of African-American ancestry from 11 different cohorts of the CHARGE consortium including: the Rotterdam Study (RS-I and RS-II, N = 4119), the Rhineland Study (N = 4019), the MEMENTO cohort (N = 2195), the Framingham Heart Study (FHS, N = 2048), the BiDirect study (N = 1899), the Cardiovascular Health Study (CHS-, African-American N = 273, European-American N = 1396), the Atherosclerosis Risk in Communities (ARIC-, African American N = 823, European American N = 742), the Vietnam Era Twin Study of Aging (VETSA, N = 828), the Alzheimer’s Disease Neuroimaging Initiative (ADNI, N = 578), the Coronary Artery Risk Development in Young Adults (CARDIA-, African-American N = 128, European-American N = 343) Study, and the Austrian Stroke Prevention Family Study (ASPS-Fam, N = 287). A detailed description of each of the participating cohorts, their genotyping information, and the quantification of NfL is described in the supplementary materials. General demographic information is provided in **Table 1**.

**Table 1:**
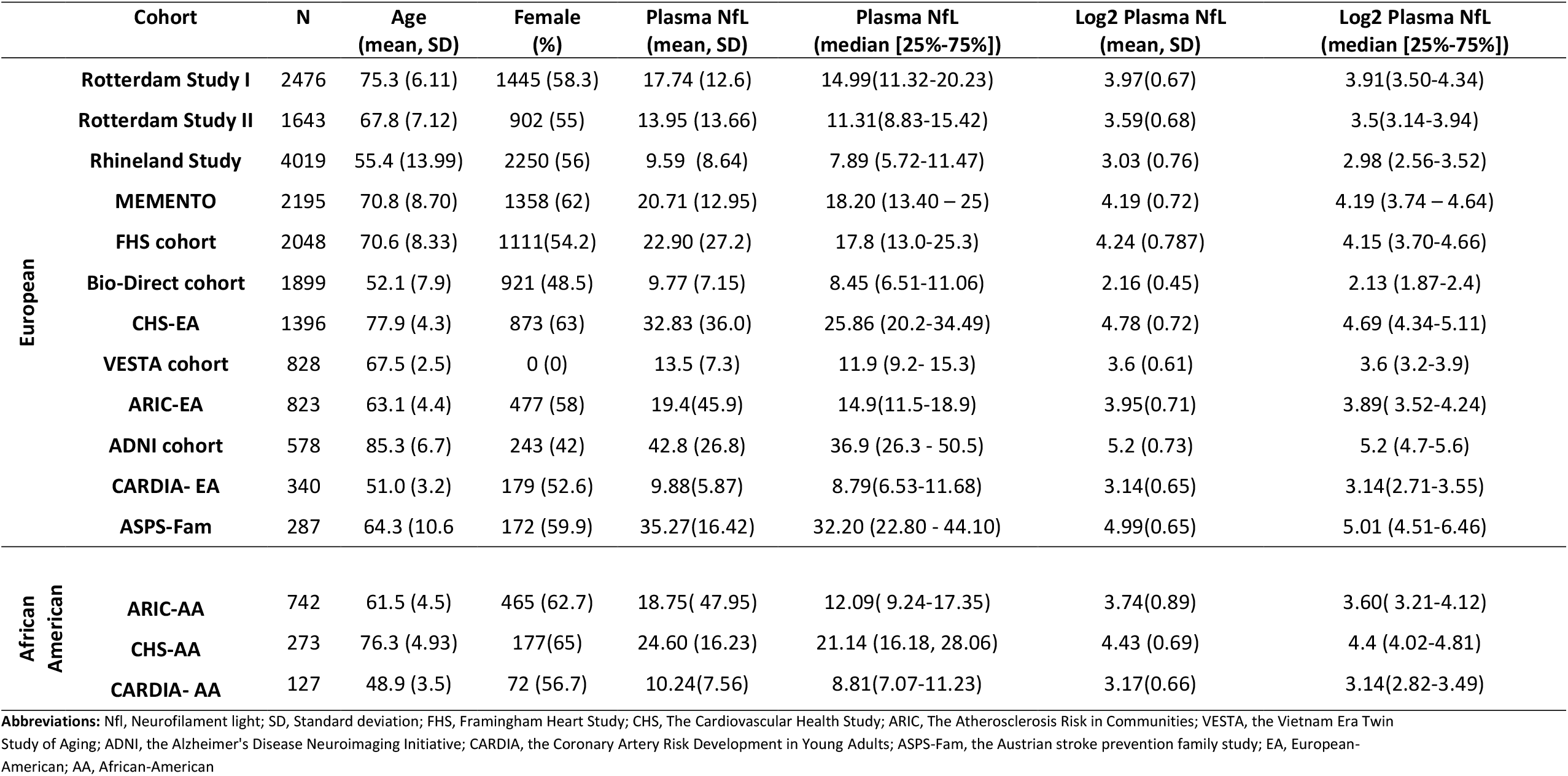
Demographic information for participating cohorts of European ancestry and African-American ancestry.

### NfL quantification

Different protocols were adopted by participating cohorts for sample preparation, plasma or serum extraction, and NfL quantification. Methodological details concerning NfL quantification are provided in the cohort descriptions included in the supplement (Supplementary table 1). In summary, the Rotterdam Study used the single molecule array (Simoa) HD-1 analyzer platform, the Rhineland study used the Quanterix Simoa NF-light assay (103186), the FHS, ARIC, and CARDIA cohorts used the Quanterix 4, the MEMENTO cohort used Simoa NF-light kit on a Quanterix H1 analyzer, BiDirect cohort profiled NFL on Simoa HDX analyzer, the CHS used the Simoa Human Neurology 4-Plex A assay and the ASPS-fam used Simoa HDX analyzer.

### Genotyping and Imputation

The participating cohorts genotyped their samples employing various genotyping kits and imputed using either 1000 Genomes (1Kg)(9) or the Haplotype Reference Consortium (HRC)(10) panels. Detailed descriptions of the genotyping and imputation methods are provided in cohort description (**Supplementary table 1)**.

### Statistical analysis

#### Ancestry specific GWAS meta-analysis

Each participating cohort performed genome-wide association of single nucleotide polymorphism (SNP) and plasma or serum levels of NfL using an additive model. Blood levels of NfL were log2 transformed before conducting the GWAS and analyses were adjusted for age, sex, study-specific covariates, and genetic principal components to account for population structure and family relatedness. A post-GWAS quality control was performed on summary statistics of each study using the easy QC software(11). We excluded SNVs with low imputation quality scores (info score or *r*^*2*^ < 0.3), low frequency (minor allele count <5 or minor allele frequency < 0.01), and variants that were available in less than 30 participants for each cohort. In order to identify ancestry-specific genetic variants, we performed an ancestry-stratified GWAS meta-analysis for three cohorts of African-American ancestry and 11 cohorts of European ancestry separately, using METAL(12) with inverse variance weighted average score to account for population heterogeneity and genomic inflation. In the European ancestry GWAS meta-analysis, we retained only 705,8703 genetic variants that were present in at least two major cohorts (i.e. the Rotterdam Study, and the Rhineland Study) of a total of 11 cohorts accounting for more than 40% of the total number of European ancestry participants. In the case of the African-American ancestry, we retained 838,1611 genetic variants present in all three cohorts of African ancestry (ARIC-AA, CHS-AA, and CARDIA-AA) due to the limited sample size. Moreover, we excluded variants with heterogeneity *I*^*2*^ values greater than 0.75 in the ancestry-specific meta-analysis.

#### Functional mapping and annotation

To perform functional mapping, and annotation of GWAS summary statistics of NfL, we used the Functional Mapping and Annotation (FUMA) platform version 1.3.8 which is designed to prioritize and aid in the interpretation of GWAS findings(13, 14). To identify independent genome-wide significant SNPs, we used *r2* = 0.2 and *P-value* < 5x10^−8^. Using FUMA, we defined the lead SNPs as independent of each other at *r2* = 0.1 within a 500 kb region in the 1000 Genome Phase 3 reference panel. The individual lead SNPs were mapped based on the default 10kb distance between SNPs and genes. The ancestry-specific GWAS meta-analysis NfL loci were visualized using Manhattan plots and regional plots using EasyStrata (15) and Locus Zoom(16) (using the 1000 Genomes reference panel for estimating LD), respectively. We used LD score (LDSC) regression software(17) to estimate blood NfL heritability based on GWAS summary statistics. Reference LD scores were computed based on the 1000 Genomes reference panel.

#### Pathway enrichment analysis and functional analysis

Gene-based and gene-set enrichment analyses which quantify the association of individual mapped genes with NfL levels and sets of genes with GO terms, respectively, were performed using MAGMA (version v1.0.8) (18) as implemented in FUMA (version 1.3.7). The gene-based analysis was performed based on 18,718 protein-coding genes, setting the level of statistical significance at a Bonferroni-adjusted threshold of *P-value* = 2.671x10^−6^ (= 0.05/18718). Similarly, tissue-specific gene expression analysis was also performed using MAGMA as integrated in FUMA. Further, we explored the effects of genetic variants identified in our GWAS on the expression levels of other genes by querying the genotype-tissue expression (GTEx)(19) database (version 8) for genes expressed in brain and blood.

#### LD regression analysis

To quantify the genetic correlation between blood NfL levels and other neurological traits and biomarkers of neurodegeneration, we performed LD regression analysis. We obtained GWAS summary statistics for AD, PD, Huntington’s disease, ALS, amyloid-beta (Aβ)-42, Aβ-40, total-tau, and brain imaging markers (total hippocampal volume, total brain volume, and total white matter lesions) from the GWAS catalogue(20). We performed LD regression analysis using the LDSC tool(17) based on the European ancestry 1000 Genomes LD reference panel. Details of the GWAS studies used for LD regression and their base heritability estimates are provided in **Supplementary table 2**.

#### Polygenic Risk Score association with AD biomarkers

We calculated genetic risk scores (GRS) based on the genome-wide significant variants leading the two independent loci of NfL in people from European ancestry. GRS was calculated in the Rotterdam Study participants by summing the number of risk alleles of rs7203642 and rs12051560 variants weighted by their regression coefficients obtained from our meta-analysis. We used the Cox proportional-hazard models to check the association of GRS with the incidence of AD, adjusted for age at baseline and sex. Moreover, we performed multiple linear regression analyses to assess the association of NfL GRS with plasma levels of Aβ-40, Aβ-42, total tau, as well as with magnetic resonance imaging markers (MRI) markers of neurodegeneration including total hippocampal volume, total brain volume, and total white matter lesions in the Rotterdam Study cohort. All linear regression analyses were adjusted for age, sex and additionally for intracranial volume for MRI traits.

#### Look up of lead variants into previous GWASs of neurological traits

To evaluate the association of the most significant genetic variants with the two common neurodegenerative diseases AD and PD, we used the most recent GWAS meta-analyses of AD(21) and PD(22) and reported the results for each genetic variant. Additionally, we performed lookups for single variants in GWASs of traits used for LD regression analysis.

## Results

Our ancestry-specific GWAS meta-analysis of circulating levels of NfL was based on 11 different cohorts of European (N = 18532) and African American ancestry (N = 1142), **Table 1**. The Rotterdam Study and the Rhineland study were the major contributors (> 40%) to the total samples size. Participants of cohorts of European ancestry had diverse age ranges, varying from a mean age of 51 years (standard deviation [SD]= 3.2) in CARDIA-EA to a mean age of 85.3 years (SD = 6.7) in the ADNI cohort. The female proportion varied from 0% in the VESTA cohort to 63% in the CHS-EA cohort. Among the three cohorts representing African-American ancestry, the ARIC-AA cohort contributed the largest number of participants, while CHS-AA participants were older (mean = 76.3 years [SD =4.93]) compared to the other two cohorts (mean ages of 61.5 [SD = 4.5] and 48.9 [SD = 3.5] years in ARIC-AA and CARDIA-AA, respectively). Moreover, Cardia-AA had the lowest percentage of female participants (56.7%). Overall mean NfL levels in all participating cohorts were significantly positively correlated with mean age (Pearson’s r = 0.81, *P* = 2.06x10^−4^).

### GWAS meta-analysis findings

The European ancestry GWAS meta-analysis identified 26 genome-wide significant SNPs within two loci lead by two individually significant SNPs (*r2* < 0.1) (**Table 2**). Manhattan plot and Quantile–Quantile (Q-Q) plot of the meta-analysis summary statistics are provided in **Figure 1:A** and **Supplementary figure 1:A**. The first locus at 16p12 was mapped to the *UMOD, PDILT* genes, and it was tagged by 39 SNPs with *P-values* < 0.05 (**Supplementary table 3**). This locus was led by two genetic variants (r2 ≥ 0.4) reaching genome-wide significance, including rs7203642-A (effect = 0.041, standard error [SE] = 0.007, *P* = 1.37x10^−8^) in the intronic region of the *UMOD* gene and rs77924615-A (effect = -0.041, SE = 0.007, *P* = 3.77x10^−8^) within the intronic region of the *PDILT* gene. A regional plot for 16p12 locus (**Figure 2:A**) shows that both of these variants are also in high LD (r2∼0.4) with each other.

**Table 2:**
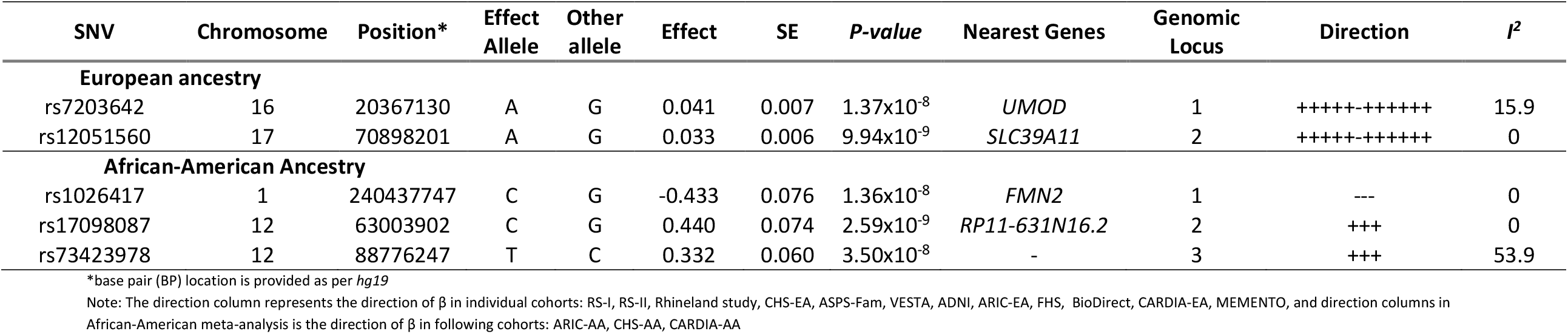
Genome-wide significant loci associated with blood levels of Neurofilament light (NfL) in European and African-American Ancestry.

**Figure 1:**
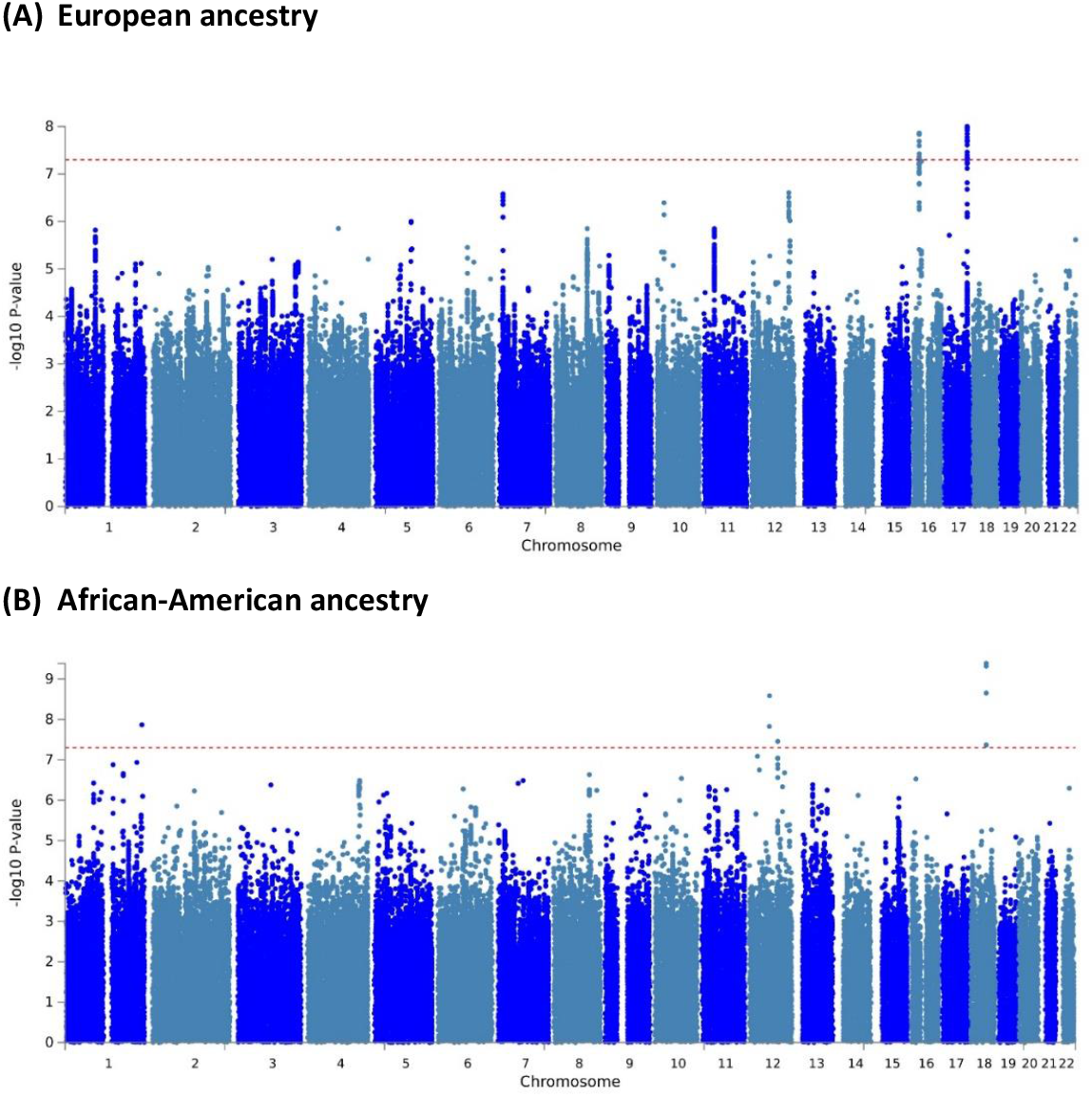
Manhattan plot for the meta-analysis of genome-wide association study (GWAS) of the blood levels of neurofilament light (NfL) in the European ancestry (**A**) and African-American ancestry (**B**). Observed association of all tested genetic variants on autosomal chromosomes (X-axis) are displayed as – log10(*P-values*) on Y-axis. Red dotted horizontal line indicates a genome-wide significant association (*P-value* <5x10^−8^) with NFL levels in blood.

**Figure 2:**
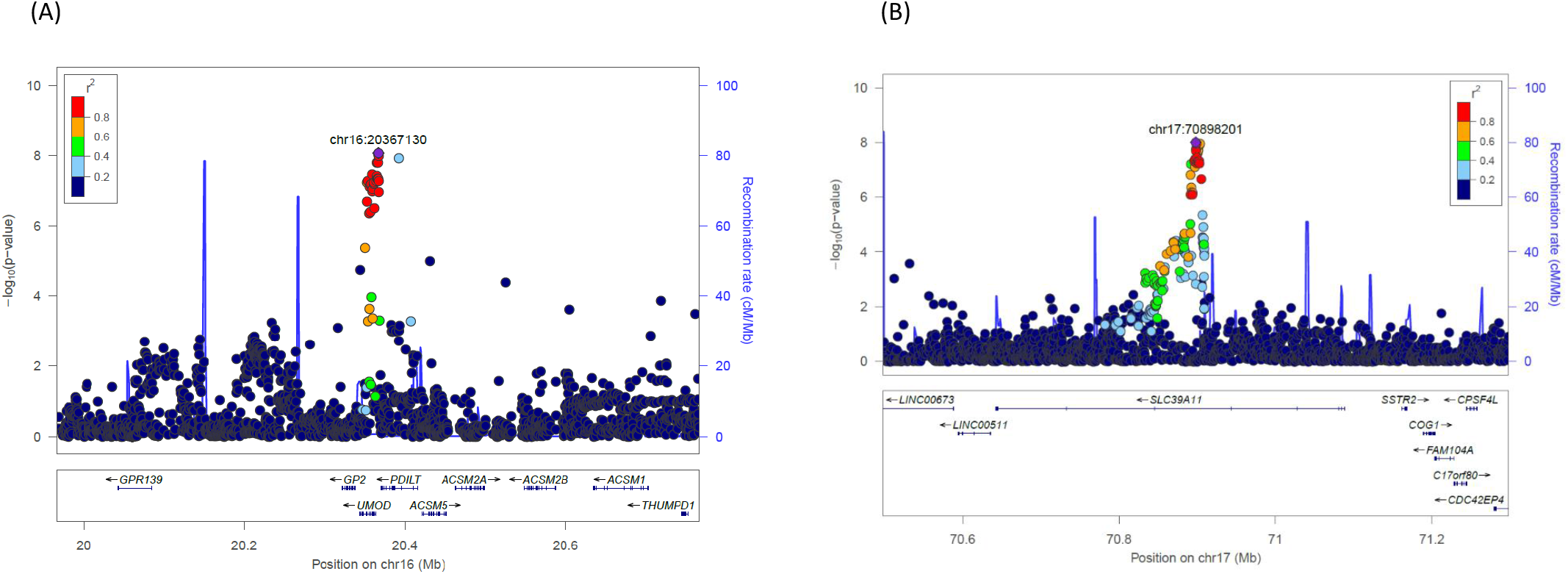
Regional plot for two loci identified in the meta-analysis of neurofilament light (NfL) genome-wide association study (GWAS) in European ancestry. The genetic variants are denoted as colored circles with their *P-values* (-log10) on left Y-axis and genomic location is based on build 37 on X-axis. Lead SNPs (purple diamond) are marked with their genomic location. Recombination rates are plotted on right Y-axis to represent the local linkage disequilibrium (LD) structure. The LD between the genetic variants is provided with a color scale, ranging from blue (r2 =0) to red (r2 =1). LD calculations are based on 1000 genome European ancestry.

**Figure 3:**
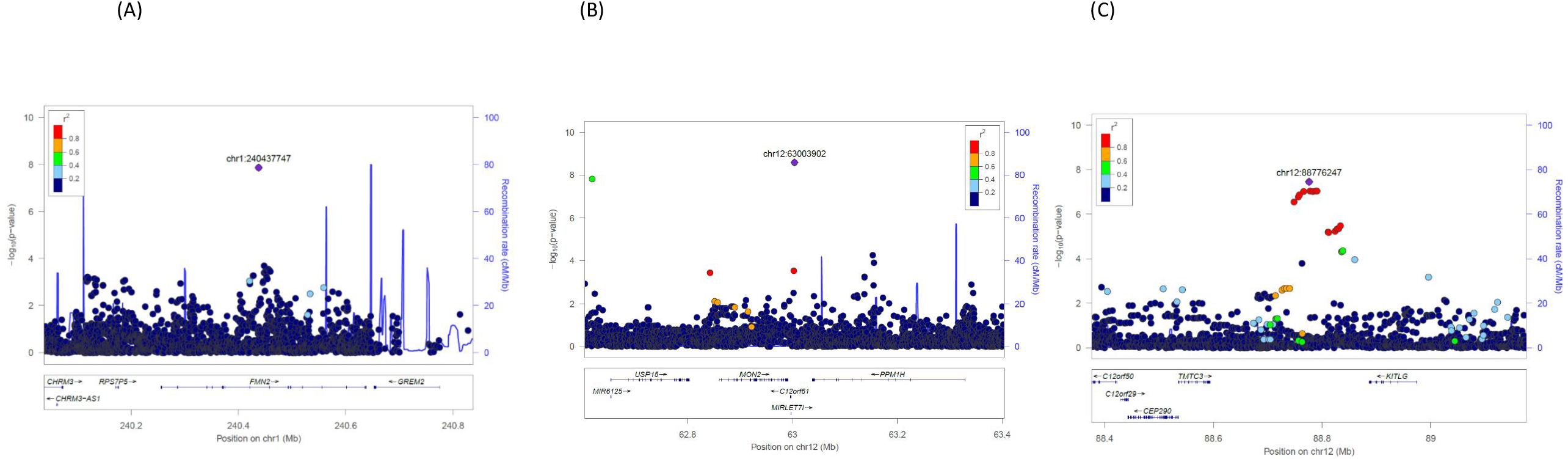
Regional plot for two loci identified in the meta-analysis of neurofilament light (NfL) genome-wide association study (GWAS) in African-American ancestry. The genetic variants are denoted as colored circles with their P-values (-log10) on the left Y-axis and genomic location is based on build 37 on X-axis. Lead SNPs (purple diamond) are marked with their genomic location. Recombination rates are plotted on right Y-axis to represent the local linkage disequilibrium (LD) structure. The LD between the genetic variants is provided with a color scale, ranging from blue (r2 =0) to red (r2 =1). LD calculations are based on 1000 genome African ancestry.

Therefore, we defined this locus based on the rs7203642 variant of *UMOD* gene, with the A-allele of rs7203642 is associated with increased blood NfL levels. The second locus at chromosome 17q24 was tagged by 117 SNPs with a *P-values* < 0.05 (**Supplementary table 3**), and was mapped to the *SLC39A11* gene. At 17q24 locus, the A-allele of the lead intronic variant rs12051560 (effect = 0.033, SE = 0.006, *P* = 9.94x10^−9^) was associated with increased blood NfL levels (**Figure 2:B** regional plot). Based on LSDC, the SNP-heritability (*h)*^*2*^ of blood NfL levels was 0.12, meaning that the identified SNPs can explain about 12% of the variation of NfL levels in blood.

The Manhattan plot and Q-Q plots for GWAS meta-analysis of African-American ancestry are provided in **Figure 1:B** and **supplementary figure 1:B**. In the GWAS meta-analysis of African-American cohorts (**Table 2**), we identified three independent genome-wide significant loci at chromosomes 1q43, 12q14, and 12q21, which were altogether tagged by 75 SNPs with *P* < 0.05 (**Supplementary Table 4**). An intronic variant inside the *FMN2* gene (rs1026417-C, effect = -0.433, SE = 0.076, *P* = 1.36x10^−8^) was associated with decreased levels of NfL in circulation, while two genetic variants at 12q14 (rs17098087-C, effect = 0.440, SE = 0.074, *P* = 2.59×10^−9^) and 12q21 (rs73423978-T, effect = 0.332, SE = 0.060, *P* = 3.50×10^−8^) were associated with increased levels of NfL in blood. We have provided the information about Combined Annotation Dependent Depletion (CADD) score, Regulome Database (RDB) annotation, and chromatin state information for all SNPs inside the observed genetic loci for both European and African-American ancestry using FUMA in **Supplementary table 3, 4** and **Supplementary figures 2-6**.

#### Conditional analysis on kidney function (eGFR)

In the European ancestry meta-analysis, the locus at 16p12 (*UMOD* gene) is also known locus for kidney function(23). Kidney function was not included as a covariate in the GWAS of blood NfL levels by the participating cohorts, although it may have a role in blood protein clearance and thus may confounding genetic associations of blood protein levels. To investigate whether genetic variants associated with kidney function may have confounded the associations between the identified SNPs and blood NfL levels, we conducted a conditional analysis by conditioning the observed genetic association effect size estimates on the estimated kidney glomerular filtration rate (eGFR)(23) associated genetic variants in European ancestry using mtCOJO(24). Results of this conditional analysis (**Supplementary table 5**) showed that one of the lead genetic variants at 17q24 (rs12051560-A, Effect = 0.033, SE = 0.006, *P* = 9.13×10^−9^) is independent of kidney function. However, the second locus inside the intronic region of *UMOD* gene became less significant (rs7203642-A, effect = 0.034, SE = 0.007, *P* = 3.51×10^−6^) upon conditioning the meta-analysis on kidney function.

#### Gene enrichment analysis

Gene enrichment analysis of both European (number of genes = 18,718) and African-American (number of genes = 17,370) ancestry-based meta-analysis showed enrichment of several GO terms, though they did not pass the Bonferroni-adjusted thresholds for multiple testing (**Supplementary table 6, 7**). We also did not find an overlap in the top ten curated GO terms in the ancestry-specific enrichment analysis. Yet, the top GO terms enriched in European ancestry meta-analysis included GO molecular function beta 2 adrenergic receptor binding (*P* = 1.89×10^−5^), GO biological process glycerolipid catabolic process (*P* = 2.97×10^−5^), germ cell proliferation (*P* = 6.85×10^−5^), and canonical wnt signaling pathway (*P* = 5.57×10^−5^).

In addition, genes were enriched in curated gene sets including sharma pilocytic astrocytoma location dn, reactome foxo mediated transcription of cell death genes, and pid p38 alpha beta downstream pathway.

In African-American ancestry meta-analysis findings were enriched for GO biological process astrocyte differentiation (*P* = 1.38×10^−4^**; Supplementary table 7**), compartment pattern specification (*P* = 1.85×10^−4^) and GO astrocyte development (*P* = 3.38×10^−4^).

#### eQTL analysis for the identified genetic variants

We then performed eQTL lookups for all the lead SNPs of identified loci in both ancestries to assess which genetic variants are associated with the expression of cis- and trans-genes. The lead genetic variant at chromosome 17, rs12051560-A allele was associated with decreased expression of the *SSTR2* gene in cerebellar hemispheres (Normalized Effect Size [NES] = - 0.28, *P* = 1.7×10^−7^) and the cerebellum (NES = -0.26, *P* = 7.2×10^−6^).

#### Genetic correlation of NfL with neurological traits

In the LD regression analysis based on the results of European ancestry meta-analysis (**Table 3**), we observed a strong positive genetic correlation of NfL with AD (r_g_ = 0.322, *P* = 1.74×10^−6^), total-tau (r_g_ = 2.014, *P* = 1.04×10^−6^), Aβ-40 (r_g_ = 0.829, *P* = 5.36×10^−6^) and Aβ-42 (r_g_ = 1.024, *P* = 6.37×10^−5^). However, r_g_ values were inflated with total-tau (*h*_*2*_ = 0.066; **Supplementary table 2**) and Aβ-42 (*h*_*2*_ = 0.092), which can be due to low heritability estimates in one of the regressed traits. We repeated the LD score regression analysis after removing *MAPT* and *APOE* region from total-tau and Aβ-42 GWAS summary statistics data, but results did not change with total-tau (rg = 2.014, *P* = 1.04×10^−6^), Aβ-40 (rg = 0.829, *P* = 5.95×10^−6^) and Aβ-42 (rg = 1.024, *P* = 6.37×10^−5^). As a sensitivity test, we also repeated the LD regression analysis using the European ancestry NfL summary statistics conditioned on kidney function (**Supplementary table 8**), but the results remained similar to the original unadjusted summary statistics.

**Table 3:**
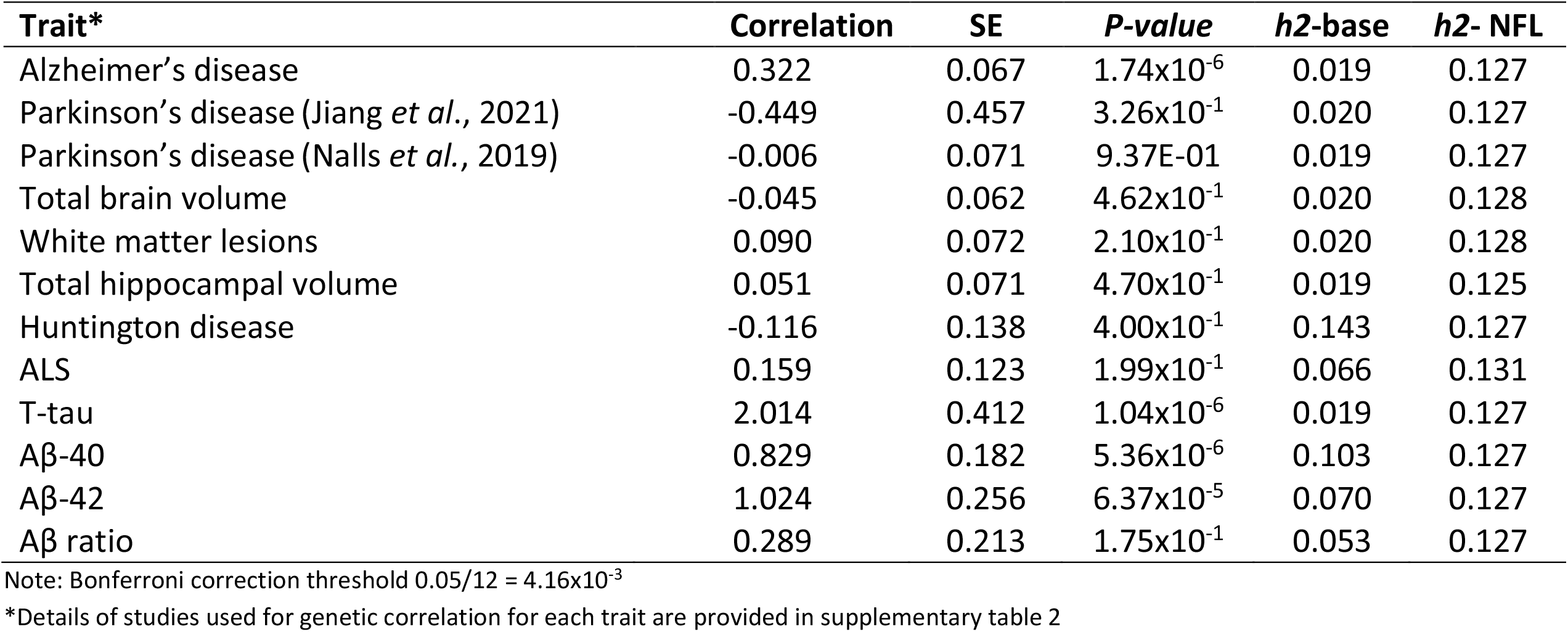
Genetic Correlation of Neurofilament light (NfL) with neurological traits.

| **Trait\*** | **Correlation** | **SE** | **P-value** | **h2-base** | **h2- NFL** |
| --- | --- | --- | --- | --- | --- |
| Alzheimer's disease | 0.322 | 0.067 | $1.74 \times 10^{-6}$ | 0.019 | 0.127 |
| Parkinson's disease (Jiang *et al.*, 2021) | -0.449 | 0.457 | $3.26 \times 10^{-1}$ | 0.020 | 0.127 |
| Parkinson's disease (Nalls *et al.*, 2019) | -0.006 | 0.071 | 9.37E-01 | 0.019 | 0.127 |
| Total brain volume | -0.045 | 0.062 | $4.62 \times 10^{-1}$ | 0.020 | 0.128 |
| White matter lesions | 0.090 | 0.072 | $2.10 \times 10^{-1}$ | 0.020 | 0.128 |
| Total hippocampal volume | 0.051 | 0.071 | $4.70 \times 10^{-1}$ | 0.019 | 0.125 |
| Huntington disease | -0.116 | 0.138 | $4.00 \times 10^{-1}$ | 0.143 | 0.127 |
| ALS | 0.159 | 0.123 | $1.99 \times 10^{-1}$ | 0.066 | 0.131 |
| T-tau | 2.014 | 0.412 | $1.04 \times 10^{-6}$ | 0.019 | 0.127 |
| Aβ-40 | 0.829 | 0.182 | $5.36 \times 10^{-6}$ | 0.103 | 0.127 |
| Aβ-42 | 1.024 | 0.256 | $6.37 \times 10^{-5}$ | 0.070 | 0.127 |
| Aβ ratio | 0.289 | 0.213 | $1.75 \times 10^{-1}$ | 0.053 | 0.127 |

Note: Bonferroni correction threshold $0.05/12 = 4.16 \times 10^{-3}$
\*Details of studies used for genetic correlation for each trait are provided in supplementary table 2

#### Genetic risk score analysis in the Rotterdam Study

We further corroborated the results of the LD regression analyses, which were based on summary statistics, by deriving a GRS based on individual-levels data (**Supplementary Table 9**). In the Rotterdam Study cohort, the GRS based on two lead genetic variants of the identified loci in European ancestry participants showed strong associations with plasma levels of total tau (β = 0.844, SE = 0.227, *P* = 1.97×10^−4^), Aβ-40 (β = 0.562, *P* = 2.24×10^−5^), and Aβ-42 (β = 0.644, *P* = 2.93×10^−4^). We did not observe associations of the NFL GRS with AD risk (β = 0.129, SE = 0.916, *P* = 8.88×10^−1^) in the Rotterdam Study.

#### Relation of identified single genetic variants with neurological traits

We also performed look-ups of the two identified genetic variants associated with blood NfL levels in European ancestry participants using recent AD(21), PD(22), and other GWAS (meta-analysis) summary statistics included in our LD regression analyses (**Supplementary table 10)**. Neither of the two genetic variants showed an association with AD or PD. Genetic variant inside the *UMOD* (rs7203642-A) was associated with all three AD biomarkers i.e. total-tau (β = 0.028, *P* = 6.21×10^−4^), Aβ-40 (β = 0.039, *P* = 1.37×10^−2^), Aβ-42 (β = 0.033, *P* = 4.43×10^−2^), while the variant inside the *SLC39A11* gene (rs12051560-A) showed nominal p-value of association with total-tau only (β = 0.012, *P* = 7.30×10^−2^).

In the association of single genetic variants of NfL with CSF levels of Aβ-42, phosphorylated tau (p-tau), total-tau in the MEMENTO cohort (**Supplementary table 11**), only one genetic variant rs12051560-A (*SLC39A11*) was nominally associated with CSF levels of total tau (β = 0.138, *P* = 4.96×10^−2^).

## Discussion

In these ancestry-specific GWAS meta-analyses, we identified two genome-wide significant loci (16p12 and 17q24) associated with blood NfL levels within the *UMOD* and *SLC39A11* genes among European ancestry participants, and three loci (1q43, 12q14, and 12q21) in participants from the African-American ancestry. Conditioning the GWAS summary statistics on kidney function indicated that the genetic variants located inside the *UMOD* gene likely exert their effect on blood NfL levels through their influence on kidney function. Evaluation of the genetic correlation of the blood NfL levels (European ancestry) with neurological traits demonstrated significant positive correlation with AD, Aβ-42, Aβ-40, and total-tau. Further, a higher NfL GRS based on genome-wide significant variants identified in participants from European ancestry was associated with plasma levels of Aβ-42, Aβ-40, and total-tau in the Rotterdam Study cohort.

Conditional analysis based on kidney function using summary statistics data discovered a novel locus at 17q24 (*SLC39A11*) that was independently associated with blood NfL levels at the genome-wide level of significance. Interestingly, there are several pieces of evidence derived from earlier studies that all point toward the role of the *SLC39A11* gene in neurodegeneration. The *SLC39A11* gene plays a role in zinc homeostasis(25) and has been associated with ALS (*P* = 8.11×10^−6^)(26, 27). The KEGG pathway database (map05010 and map05012) queries indicated a role of the *SLC39A11* gene in the AD-as well as PD-related pathways(28). Moreover, an *RNA* expression-based study in AD and Huntington’s disease identified the *SLC39A11* gene as a common hub gene of the differentially expressed genes(29). Another interesting observation that links the *SLC39A11* polymorphisms to neuro-axonal injury is that the rs12051560-A (*SLC39A11*) associated with decreased expression of the somatostatin receptor 2 (*SSTR2*) gene in the cerebellum and the cerebellar hemispheres based on the queries of the GTEx eQTL database. Decreased expression of *SSTR2* gene was linked to axonal degeneration of noradrenergic projections in SSTR2-/- mice studies(30). The *SSTR2* gene plays a role in the integrity and maintenance of the noradrenergic system, a brain network involved in the regulation of memory, stress, emotions, motor coordination, and arousal functions(31). Moreover, evidence from earlier studies indicated the involvement of the *SSTR2* gene in neurodegeneration under ischemia(32) and G_s_ receptors coupled cAMP level reduction (33), which plays a crucial role in neuronal survival and differentiation in cell culture conditions(34). Earlier studies also demonstrated the role of somatostatins, substrate of somatostatin receptors, in hypoxia-induced neuronal cell death(35) due to their involvement in potassium channel activation(36).

Another locus for blood NfL levels in European ancestry participants was identified in the 16p12.3 region, which is tagged by two lead genetic variants located inside the *UMOD* and *PDILT* genes, which have previously been associated with kidney function (23). Conditional analysis on the kidney function showed that the association of our lead SNP rs7203642 became less significant (*P* = 3.51×10^−6^). The association of blood NfL levels with variants in *UMOD* is supported by results from earlier studies reporting the association between increased blood NfL levels and decreased kidney function, which may be due to aging, cardiovascular risk factors, and diabetes mellitus (37-40). The observation of higher blood levels of NfL in children with chronic kidney disease (41) contradicts the role of cardiovascular diseases and diabetes as the sole drivers of blood NfL levels. Nevertheless, kidney function has been associated with cognitive decline, brain atrophy, and white matter abnormalities(42, 43) which suggest a direct role of kidney function in determining blood NfL levels. The exact mechanism through which genetic variants in the *UMOD* and blood levels of NfL are related needs further investigation, but the key role of *UMOD* in kidney function may be instrumental in understanding its link to neurodegeneration. The *UMOD* gene encodes for uromodulin protein and mutations in this gene have been associated with hyperuricemia and tubulointerstitial nephritis(44). One of the primary reasons for *UMOD*-related kidney disease is the accumulation of misfolded *UMOD* protein inside the endoplasmic reticulum (ER)(45) and thereby generation of ER stress which results in cell death and inflammation(46). Hyperglycemia and ER stress related pathways may trigger the generation of advanced glycation end products (AGE), which have been associated with neurodegeneration(46, 47). Second, the lead genetic variants of identified locus i.e., rs77924615 was found inside the *PDILT* gene, which belongs to the protein-disulfide isomerase (PDI) family of proteins(48). PDI proteins play an important role in protein folding in the ER and their dysfunction may lead to diseases involving the accumulation of misfolded proteins, which is also a hallmark of neurodegenerative diseases such as AD and PD(23).

Genetic correlation of the European ancestry-based NfL summary statistics with other neurological traits demonstrated a strong genetic correlations of NfL with AD, and AD-related biomarkers including blood-based t-tau, Aβ-40, and Aβ-42. The higher genetic correlation between NfL and AD biomarkers could partly be attributed to kidney function loci/locus, as kidney function could influence the clearance of protein-based biomarkers in blood. Indeed, genetic variants inside the *UMOD* gene also showed nominally significant associations (*P* < 0.05; **Supplementary table 10**) with all three biomarkers in previously published GWAS. However, our sensitivity analysis, using LD regression analysis with eGFR-adjusted (conditional analysis) NfL summary statistics produced similar results, indicating that the genetic correlation of NfL with biomarkers of neurodegeneration is largely independent of kidney function. This conclusion is further supported by our findings of strong associations of NfL GRS with t-tau, Aβ-40 and Aβ-42 levels in plasma in the Rotterdam Study, though we did not observe an association with incident AD which maybe due to our relatively small number of incident AD cases. Further, variants inside the *SLC39A11* gene were also associated with higher levels of total tau in CSF (*P*<0.05) based on our post-GWAS analysis in the MEMENTO cohort, supporting the notion that genetic determinants of blood NfL levels are likely linked to central neurodegeneration.

In the participants from the African-American ancestry, one of the three loci was located near the *FMN2* gene (rs1026417-C). *FMN2* is a coding gene involved in the cytoskeleton assembly, which makes it an interesting discovery since NfL is a cytoskeleton protein released into extracellular space as a result of neuro-axonal damage(5). *FMN2* gene is highly expressed in the brain and involved in synaptic plasticity and memory formation(49). Our observation was concordant with several studies that reported the association of the *FMN2* gene with cognition(50), ALS (51), intellectual development disorder(52) and neuropsychiatry traits(53). All these interlinked evidences support the role of the *FMN2* gene in determining the blood NfL levels in various neurological diseases in African-American ancestry. None of the genetic variants identified in the African-American ancestry meta-analysis showed association with blood NfL levels in the European ancestry.

Next, in the pathway-enrichment analysis based on the European ancestry, we did not identify significant enrichment of GO biological processes for our observed genes after multiple testing correction. However, GO: canonical wnt signaling pathway (*P* = 5.57×10^−5^) and GO: regulation of wnt signaling pathway (*P* = 1.43×10^−4^) were among the top pathways that were enriched for 307 and 334, respectively, of the putative genes identified in our study. Wnt signaling is one of the most crucial pathways involved in brain development and involves several genes associated with neurodegenerative diseases such as AD and PD(54). Furthermore, the ‘beta 2 adrenergic receptor’ binding GO molecular process ranked first in our analysis (*P* = 1.89×10^−5^). Interestingly, blocking the beta 2 adrenergic receptors is found to be an effective approach in PD to reduce neuroinflammation and degeneration of dopaminergic neurons(55, 56). In the African-American ancestry, in GO biological processes, go astrocyte differentiation (*P* = 1.38×10^−4^) and go astrocyte development (*P* = 3.38×10^−4^) were most notable terms enriched in the GWAS, which reiterates the role of astrocytes in neurodegeneration(57).

Our study represents the first largest GWAS to uncover the genetic determinants of NfL levels in blood. Our GWAS sample included 11 different cohorts of both European and African-American ancestry which is the main strength of our study. Two genetic variants identified in our study not only highlight the importance of kidney function in neurodegeneration but also indicate that the kidney function should be taken into account when assessing blood-based protein biomarkers and specifically NfL. This study has also limitations. A small sample for African-American cohorts was a major limitation for performing a trans-ethnic meta-analysis.

In conclusion, we identified two unique loci associated with blood levels of NfL in participants from European ancestry and three loci in African-American ancestry participants. The genetic locus inside *SLC39A11* gene represents a promising candidate that could be involved in a common pathways underlying axonal damage and neurodegeneration in European ancestry. Moreover, our findings highlight the role of the *UMOD* and *PDILT* genes, which are involved in protein miss-folding and renal accumulation of uromodulin, in linking reduced kidney function to neuro-axonal injury and neurodegeneration

## Supporting information

Supplementary Methods Description

## Data Availability

All data produced in the present study are available upon reasonable request to the authors and subject to legal and ethical approvals.

## Acknowledgements

**Rotterdam Study cohort:** The Rotterdam Study is supported by the Erasmus MC University Medical Center and Erasmus University Rotterdam, the Netherlands Organization for Scientific Research (NWO), the Netherlands Organization for Health Research and Development (ZonMW), the Research Institute for Diseases in the Elderly (RIDE), the Ministry of Education, Culture and Science, the Ministry of Health, Welfare and Sport, The European Commission (DGXII), the Netherlands Genomics Initiative (NGI), and the Municipality of Rotterdam. Cardiovascular Health Study (CHS) cohorts: This CHS research was supported by NHLBI contracts HHSN268201200036C, HSN268200800007C, HHSN268201800001C, N01HC55222, N01HC85079, N01HC85080, N01HC85081, N01HC85082, N01HC85083, N01HC85086;, HHSN268200960009C and NHLBI grants U01HL080295, R01HL087652, R01HL105756, R01HL103612, R01HL120393, and U01HL130114 with additional contribution from the National Institute of Neurological Disorders and Stroke (NINDS). Additional support was provided through R01AG023629, R01AG033193, R01AG053325, and K24AG065525 from the National Institute on Aging (NIA). A full list of principal CHS investigators and institutions can be found at CHS-NHLBI.org. The provision of genotyping data was supported in part by the National Center for Advancing Translational Sciences, CTSI grant UL1TR001881, and the National Institute of Diabetes and Digestive and Kidney Disease Diabetes Research Center (DRC) grant DK063491 to the Southern California Diabetes Endocrinology Research Center. The content is solely the responsibility of the authors and does not necessarily represent the official views of the National Institutes of Health. **The BiDirect Study:** The BiDirect Study is supported by grants (01ER0816, 01ER1506) of the German Ministry of Research and Education (BMBF) to the University of Muenster. Laboratory NFL analysis was performed at the University Hospital Basel and in addition supported by a grant of the Swiss National Science Foundation. **The Framingham Heart Study (FHS)**: This work was supported by the National Heart, Lung and Blood Institute’s Framingham Heart Study Contract No. N01-HC-25195 and No. HHSN268201500001I, and by grants from the National Institute of Aging (R01s AG033193, AG008122, AG054076, AG033040, AG049607, AG05U01-AG049505), and the National Heart, Lung and Blood Institute (R01 HL093029, HL096917). The laboratory work for this investigation was funded by the Division of Intramural Research, National Heart, Lung, and Blood Institute, National Institutes of Health, and by NIH contract N01-HC-25195. The analytical component of this project was funded by the Division of Intramural Research, National Heart, Lung, and Blood Institute, and the Center for Information Technology, National Institutes of Health: **MEMENTO cohort :** The MEMENTO cohort was sponsored by the Fondation Plan Alzheimer (Alzheimer Plan 2008–2012). This work was also supported by the following: CIC 1401-EC, Bordeaux University Hospital, Inserm, and the University of Bordeaux. **The VETSA study** was supported by Grants R03 AG065643, R01 AG050595, and R01 AG076838, K01 AG063805, and K24 AG046373 from the National Institute on Aging. The content of this manuscript is solely the responsibility of the authors and does not necessarily represent the official views of the NIA/NIH, or the VA. The U.S. Department of Veterans Affairs has provided financial support for the development and maintenance of the Vietnam Era Twin (VET) Registry. Numerous organizations have provided invaluable assistance in the conduct of the VET Registry, including: Department of Defense; National Personnel Records Center, National Archives and Records Administration; Internal Revenue Service; National Opinion Research Center; National Research Council, National Academy of Sciences; the Institute for Survey Research, Temple University. This material was, in part, the result of work supported with resources of the VA San Diego Center of Excellence for Stress and Mental Health Healthcare System. Most importantly, the VETSA co-authors gratefully acknowledge the continued cooperation and participation of the members of the VET Registry and their families as well as the contributions of many staff members and students. **ASPS-Fam:** The Medical University of Graz and the Steiermärkische Krankenanstaltengesellschaft support the databank of the ASPS-Fam. The research reported in this article was funded by the Austrian Science Fund (FWF) grant numbers PI904, P20545-P05 and P13180 and supported by the Austrian National Bank Anniversary Fund, P15435 and the Austrian Ministry of Science under the aegis of the EU Joint Programme-Neurodegenerative Disease Research (JPND)-www.jpnd.eu. **ADNI:** Data used in preparation of this article were obtained from the Alzheimer’s Disease Neuroimaging Initiative (ADNI) database (adni.loni.usc.edu). As such, the investigators within the ADNI contributed to the design and implementation of ADNI and/or provided data but did not participate in analysis or writing of this report. A complete listing of ADNI investigators can be found at: <http://adni.loni.usc.edu/wp-content/uploads/how_to_apply/ADNI_Acknowledgement_List.pdf>.

## Notes

### Competing Interest Statement

The authors have declared no competing interest.

### Funding Statement

Funding sources are mentioned in the manuscript

