## Supplementary Methods Description for "Genome-wide Association Study Meta-analysis of Neurofilament light (NfL) levels in blood reveals novel loci related to neurodegeneration"

### Author's List:


Neuro-CHARGE Working group:

Shahzad Ahmad*[1,2], Mohammad Aslam Imtiaz*[3], Aniket Mishra[6], Ruiqi Wang[7], Marisol Herrera-Rivero[8,9], Joshua C Bis[10], Myriam Fornage[11], Gennady Roshchupkin[1], Edith Hofer[12,13], Mark Logue[14,15], WT Longstreth Jr[16], Rui Xia[11], Vincent Bouteloup[6], Thomas Mosley[17], Lenore Launer[18], Michael Khalil[19] , Jens Kuhle[20], Robert A. Rissman[21] ,Genevieve Chene[6], Carole Dufouil[6], Luc Djoussé[22], Michael J. Lyons[23], Kenneth J. Mukamal [24], William S. Kremen[24], Carol E. Franz[25], Reinhold Schmidt[12], Stephanie Debette[6], Monique M.B. Breteler[3,5], Klaus Berger[26], Qiong Yang[7], Sudha Seshadri[7], N. Ahmad Aziz[3,4], #, Mohsen Ghanbari[1], #, M. Arfan Ikram[1], #

Author's Affiliations:

[1] Erasmus University Medical Center, Rotterdam, the Netherlands.
[2] Division of Systems Biomedicine and Pharmacology, Leiden Academic Centre for Drug Research, Leiden University, Leiden, the Netherlands.
[3] Population Health Sciences, German Center for Neurodegenerative Diseases (DZNE), Bonn, Germany.
[4] Department of Neurology, Faculty of Medicine, University of Bonn, Bonn, Germany.
[5] Institute for Medical Biometry, Informatics and Epidemiology (IMBIE), Faculty of Medicine, University of Bonn, Bonn, Germany.
[6] University of Bordeaux, Inserm UMR 1219, Bordeaux, France.
[7] Boston University, Boston, MA.
[8] Department of Genetic Epidemiology, Institute of Human Genetics, University of Münster, Münster, Germany.
[9] Department of Psychiatry, University of Münster, Münster, Germany.
[10] Cardiovascular Health Research Unit, Department of Medicine, University of Washington, Seattle, WA, USA.
[11] Brown Foundation Institute of Molecular Medicine, McGovern Medical School, University of Texas Health Science Center at Houston, Houston, TX, USA.
[12] Clinical Division of Neurogeriatrics, Department of Neurology, Medical University of Graz, Austria.

[13] Institute for Medical Informatics, Statistics and Documentation, Medical University of Graz, Austria.
[14] National Center for PTSD, Behavioral Sciences Division at VA Boston Healthcare System, Boston, MA, USA.
[15] Department of Psychiatry and Biomedical Genetics, Boston University School fo Medicine, Boston, MA, USA.
[16] Departments of Neurology and Epidemiology, University of Washington, Seattle, WA, USA.
[17] MIND Center, University of Mississippi Medical Center, Jackson, MS, USA.
[18] Laboratory of Epidemiology and Population Science, NIH Intramural Research Program, Bethesda, MD.
[19] Department of Neurology, Medical University of Graz, Austria.
[20] Research Center for Clinical Neuroimmunology and Neuroscience University Hospital Basel Switzerland.
[21] Department of Neurosciences, University of California, San Diego, La Jolla, CA, USA.
[22] Brigham and Women's Hospital, Harvard Medical School, Boston, MA.
[23] Department of Psychological & Brain Sciences, Boston University, Boston, MA, USA.
[24] Beth Israel Deaconess Medical Center, Harvard Medical School, Boston, MA.
[25] Department of Psychiatry and Center for Behavior Genetics of Aging, University of California, San Diego, La Jolla, CA, USA.
[26] Institute of Epidemiology and Social Medicine, University of Münster, Münster, Germany.

**Cohort Description:**

**The Alzheimer's Disease Neuroimaging Initiative (ADNI) Study**

The ADNI was launched in 2003 as a public-private partnership, led by Principal Investigator Michael W. Weiner, MD. ADNI is a global research study that actively supports the investigation and development of treatments that slow or stop the progression of Alzheimer's disease. In this multisite longitudinal study, researchers at 63 sites in the US and Canada track the progression of AD in the human brain with clinical, imaging, genetic, and biospecimen biomarkers through the process of normal aging, early mild cognitive impairment (EMCI), and late mild cognitive impairment (LMCI) to dementia or AD. The overall goal of ADNI is to validate biomarkers for use in Alzheimer's disease clinical treatment trials. The primary goal of ADNI has been to test whether serial magnetic resonance imaging (MRI), positron emission tomography (PET), other biological markers, and clinical and neuropsychological assessment can be combined to measure the progression of mild cognitive impairment (MCI) and early Alzheimer's disease (AD). For up-to-date information, see www.adni-info.org.

Data used in preparation of this article were obtained from the Alzheimer's Disease Neuroimaging Initiative (ADNI) database (adni.loni.usc.edu). Phenotypic and genetic data were downloaded from the data repository hosted at the Laboratory of Neuroimaging (LONI) at the University of Southern California, the LONI Image & Data Archive (IDA). Principal Component Analyses (PCA) were performed using Eigensoft based on pruned genetic data, with exclusion of complex and HLA regions. Ethnic outliers were removed based on 6SD from the mean. Plasma tau was analyzed with the Human Total Tau kit (research use only grade, Quanterix, Lexington, MA) on the Simoa HD-1 analyzer (CE marker). ADNI1 SNP genotype data were used to perform the GWAS (Illumina Human610-Quad BeadChip). Only non-Hispanic whites ADNI1 participants were included in the GWAS of circulating tau levels. Winsorizing at 4 SD was used to removed outliers. QC on genetic data was performed (call-rate > 0.99, P-value of HWE > 10-4; MAF > 1%). Calculation of an empirical kinship matrix was performed to account for relatedness in the association analyses. Linear mixed-effects models were used to evaluate the association of genetic variants with circulating total-tau levels, adjusted for age, sex, and PCs.

**The Rotterdam Study (RS1 and RS2)**

The Rotterdam Study is a population-based cohort study among inhabitants of a district of Rotterdam (Ommoord), the Netherlands, that aims to examine the determinants of disease and health in the elderly with a focus on neurogeriatric, cardiovascular, bone, and eye disease.[8] In 1990-1993, 7,983 persons aged ≥ 55 years participated and were re-examined every 3 to 4 years (Rotterdam Study I). In 2000-2001, the cohort was expanded by 3,011 persons who were of the same age but had not yet been part of the Rotterdam Study (Rotterdam Study II) and recently moved into the area. All participants had blood collected during their first center visit, which was followed by DNA extraction.

***Genotyping and imputations:*** Genotyping was done in participants with high-quality extracted DNA in 2007-2008 and was performed at the Human Genotyping Facility, Genetic Laboratory Department of Internal Medicine, Erasmus MC, Rotterdam, The Netherlands. Imputation of SNPs was established using the Michigan Imputation server and the HRC reference panel. More specifically, the SHAPEIT2 software was used (v2.r790) to phase the data and Minimac 3 was employed for imputation to the HRC reference panel (v1.0). QC included deletion of participants with a genotype completion rate (<90%), a low genotype call rate (<95%), sex-mismatches, duplicate pairs (just one participant), uncalled variants in over 5% of the individuals and significant violations of the expected Hardy–Weinberg Equilibrium proportions (P<10−6).

***Aβ, total-tau and NfL Assessment of plasma:*** Blood was sampled in EDTA treated containers and centrifuged. Subsequently plasma was aliquoted and frozen at -80$^{0}$C according to standard procedures. Measurements were done in two separate batches. The first batch included in total 2,000 samples, obtained from a random selection of 1,000 participants from sub-cohort RS-I-4 and 1,000 from RS-II-2. The second batch included in total 3,094 samples from the remaining participants. All measurements were performed at Quanterix (Lexington, MA, USA) on a single molecule array (Simoa) HD-1 analyzer platform.[1] Samples were tested in duplicate. Two quality control (QC) samples were run on each plate for each analyte. NfL was measured with the NF-light advantage kit.[2] The Simoa Human Neurology 3-Plex A assay (N3PA) was used for measuring the concentration of total-tau, Aβ-42 and Aβ-40. When duplicates or single measurements were missing or in the case the concentration coefficient of variation (CV) exceeded 20% or control samples were out of range, participant's data were excluded from the analyses[3].

**The Rhineland Study**

The Rhineland Study is an ongoing single-center, population-based prospective cohort study among people aged 30 years and above in Bonn, Germany. All individuals living in two pre-defined recruitment areas are invited to participate in the study, and they are predominantly German of Caucasian descent. The only exclusion criterion is an insufficient command of the German language to provide informed consent. A primary objective of the Rhineland Study is to identify determinants and markers of healthy and unhealthy aging. The study uses a deep-phenotyping approach and has a special focus on brain aging. At baseline, participants complete an 8-hour in-depth multi-domain phenotypic assessment and various types of biomaterials (blood, urine, stool, and hair) are collected. Approval to undertake the study was obtained from the ethics committee of the University of Bonn, Medical Faculty. We obtained written informed consent from all participants in accordance with the Declaration of Helsinki.

***NFL quantification***: Blood samples were collected in Vacutainer EDTA tubes and centrifuged at 2000 x g for 10 min at room temperature. The obtained plasma samples were aliquoted and stored at -80 °C. NfL levels were assessed using the Simoa® NF-light Kit (103186) in a HD-1 Analyzer (Quanterix, Billerica, USA), following manufacturer's instructions. Briefly, plasma samples were thawed with the help of the 96 FluidX Tube Rack Thawing Station (Brooks Life Sciences, Darmstadt, GE) and centrifuged at 3000 x g for 10 min at 4 °C. On-board the instrument, prepared samples were diluted using the standard 4x dilution protocol and incubated with target antibody coated paramagnetic beads and biotinylated detector antibody, followed by addition of streptavidin-ß-galactosidase enzyme complex. If NfL has been captured and labeled on the bead, the enzyme hydrolyzes the resorufin β-D-galactopyranoside substrate into a fluorescent product, which provides the signal for measurement. The NfL concentrations in pg/mL were assessed from the calibration curve generated for each assay. Intra and inter-assay coefficients of variation for plasma controls were lower than 20%, in accordance with manufacturer's recommendation.

***Genotyping:*** DNA extracted from buffy coat samples were genotyped using Infinium Omni2.5 Exome-8 BeadChip containing 2,612,357 SNPs and processed using GenomeStudio (version 2.0.5). Quality control of genotypes was performed using PLINK software (version 1.9). We used 1000 Genomes phase 3 reference panel using hg19 version for imputation of missing genotypes using impute2 software. GWAS association methods was performed using rvtest software and using standard score test.

**The Cardiovascular Health Study (CHS) cohort**

The Cardiovascular Health Study (CHS) is a population-based cohort study of risk factors for coronary heart disease and stroke in adults ≥65 years conducted across four field centers [PMID: 1669507]. The original predominantly European ancestry cohort of 5,201 persons was recruited in 1989-1990 from random samples of the Medicare eligibility lists; subsequently, an additional predominantly African-American cohort of 687 persons was enrolled for a total sample of 5,888. Blood samples were drawn from all participants at their baseline examination and DNA was subsequently extracted from available samples. CHS was approved by institutional review committees at each field center and individuals in the present analysis had available DNA and gave informed consent including consent to use of genetic information for the study of cardiovascular disease. For this ancillary study, all participants who underwent routine oral glucose tolerance testing at the 1996-1997 clinic visit were included. Entry criteria for the OGTT included in-person attendance in 1996-1997, fasting status, and absence of anti-diabetic medication.

***NFL quantification:*** Serum NfL was measured in singlet with the Quanterix single molecule array platform at the CHS Central Laboratory at the University of Vermont using the HD-X analyzer and the Simoa Human Neurology 4-Plex A assay. Preliminary studies on ~200 duplicate samples demonstrated very high reproducibility. The detectable range was 0.416 - 1648 pg/mL. Inter-assay coefficients of variation ranged from 8.53 – 10.68%.

***Genotyping:*** Genotyping was performed at the General Clinical Research Center's Phenotyping/Genotyping Laboratory at Cedars-Sinai among CHS participants who consented to genetic testing and had DNA available using the Illumina 370CNV BeadChip system (for European ancestry participants, in 2007) or the Illumina HumanOmni1-Quad_v1 BeadChip system (for African-American participants, in 2010). European ancestry participants were excluded from the GWAS study sample due to the presence at study baseline of coronary heart disease, congestive heart failure, peripheral vascular disease, valvular heart disease, stroke or transient ischemic attack or lack of available DNA. Beyond laboratory genotyping failures, participants were excluded if they had a call rate<=95% or if their genotype was discordant with known sex or prior genotyping (to identify possible sample swaps). After quality control, genotyping was successful for 3,268 European ancestry and 823 African-American participants.

SNP exclusions  In CHS, the following exclusions were applied to identify a final set of 306,655 autosomal SNPs: call rate < 97%, HWE P < 10-5, > 2 duplicate errors or Mendelian inconsistencies (for reference CEPH trios), heterozygote frequency = 0, SNP not found in HapMap. Imputation to the HRC r1.1 2016 panel was performed on the Michigan imputation server. SNPs were excluded for variance on the allele dosage ≤0.01.

**The BiDirect cohort**

The BiDirect Study was initiated in 2009 as a prospective, observational study integrating three cohorts: 1) community-dwelling adults (control diagnostic group), 2) patients with an acute depressive episode (depression diagnostic group), and 3) patients who recently suffered from acute myocardial infarction (MI diagnostic group). The study, whose principal goal is the exploration of the bidirectional relationship between depression and subclinical arteriosclerosis, recruited participants in the district of Münster, Germany, and carried out extensive phenotyping and follow-up of all cohorts in parallel. The study design and methods have been described in detail in Teismann et al., 2014[4]. All participants provided informed written consent. The BiDirect Study was approved by the ethics committee of the University of Münster and the Westphalian Chamber of Physicians in Münster, North-Rhine-Westphalia, Germany, and is funded by the German Federal Ministry of Education and Research (grant numbers FKZ-01ER0816 and -01ER1506).

**NFL quantification:** Quantification of serum NfL was conducted at the University Hospital Basel, Switzerland, using the single molecule array (Simoa®) HDX analyzer (Quanterix, Lexington, MA, USA).  Measurements of serum NfL were obtained from non-fasting blood samples collected at the first visit, using the Simoa® NF-light Advantage Kit. The sNfL values obtained at initial assessment were log2-transformed and used for analysis.

**Genotyping:** Genomic DNA was isolated from whole blood samples with EDTA using standard DNA extraction kits and procedures at the University of Münster. Whole-genome genotyping was performed with the Infinium PsychArray BeadChip v1 (Illumina) at Life&Brain GmbH (Bonn, Germany). Following basic quality control of the genotyped data (HWE p>10-6, MAF>=1%, SNP and sample call rate >=95%), imputation was performed with SHAPEIT and IMPUTE2 using the 1000 Genomes Project, phase 3, European population reference panel. Individuals were removed from the sample based on relatedness, outlier detection and missing serum NfL data. Genomic coordinates are given in the hg19 genome build. Association analysis method: An additive linear regression model adjusting for Age, Sex, Diagnostic group (depression/MI/control) and first 20 Principal Components was applied using Plink 1.9.

**The Framingham Heart Study (FHS)**

The FHS is a single-site, community-based, ongoing cohort study that was initiated in 1948 to investigate prospectively the risk factors for CVD. The Original cohort (n=5209) were randomly recruited in the town of Framingham, MA, USA, and examined every 2 year since 1948; their children and spouses of the children (n=5124), the Offspring cohort, were enrolled in 1971 and examined approximately every 4 year since 1971. This study includes 2048 Offspring subjects. Written informed consent to genetic research has been obtained on all individuals included in this study. Ethics permission for FHS and genetic research in FHS was obtained from the Institutional Review Board of Boston University Medical Campus (IRB number H-32132, H-26671).

***NFL quantification:*** NFL levels were quantified using the the Quanterix 4 plex and assays at University of Vermont by Russ Tracy.

***Genotyping**:* Genotyping was performed on Affymetrix 500K mapping array plus Affymetrix 50K supplemental array. Pre-imputation QC filter include excluding genotyped SNPs call rate<97%, HWE P<10-6, MAF<1%, Number of Mendelian errors>=1000, or at locations that did not map to GRCh37. Genotyping data was imputed using Haplotype Reference Consortium panel (hg19) using university of Michigan imputation server. Association analysis was performed using R software, lmekin function.

**The Austrian stroke prevention family study (ASPS-Fam)**

The Austrian stroke prevention family study (ASPS-Fam) is a prospective single-center, community-based study on the cerebral effects of vascular risk factors in the normal elderly population of the city of Graz, Austria. The ASPS-Fam represents an extension of the Austrian stroke prevention study (ASPS), which was established in 1991[5,6]. Between 2006 and 2013, study participants of the ASPS and their first grade relatives were invited  to enter ASPS-Fam. Inclusion criteria were no history of previous stroke or dementia and a normal neurologic examination. The entire cohort underwent an extended diagnostic work-up including clinical history, blood tests, cognitive testing, MR imaging and a thorough vascular risk factor assessment. A total of 419 individuals were included into the study and 287 participants from 153 families had GWAS and NFL data. The study protocol was approved by the ethics committee of the Medical University of Graz, Austria, and written informed consent was obtained from all subjects[7,8].

***NFL quantifications:*** All serum samples were analyzed at the University Hospital Basel, Switzerland. Serum NfL levels were determined by single molecule array (Simoa) assay using the capture monoclonal antibody (mAB) 47:3 (initial dilution 0.3 mg/mL; Art. No. 27016) and the biotinylated detector mAB 2:1 (0.1 µg/mL; Art. No. 27018) from Uman Diagnostic[9] transferred onto the Simoa platform[10]. The samples from the same participants were analyzed together in the same run to avoid within-subjects run-to-run variability. Intra- and interassay variability of the measurements were evaluated with three native serum samples in five consecutive runs on independent days. The mean coefficients of variation (CVs) of duplicate determinations for concentration were 8.5% (9.5 pg mL-1, sample 1), 5.4% (23.2 pg mL-1, sample 2) and 7.8% (98.5 pg mL-1, sample 3). Interassay CVs for serum were 7.8% (sample 1), 8.3% (sample 2) and 4.9% (sample 3)[11].

***Genotyping*:** Genotyping was performed using the Affymetrix Genome-Wide Human SNP Array 6.0. Genotypes were imputed using HRC (r1.1 2016) imputation panel using Michigan Imputation Server. Pre-imputation quality control include sample call rate 98%, SNP call rate 98%, MAF

0.05, HWE p-value < 5*10-6. Other sample exclusion criteria include sample failures, sex mismatch, cryptic relatedness, heterozygosity: mean+-3SD.

**The MEMENTO cohort**

The MEMENTO study prospectively included 2,323 individuals of French memory clinic, who presented either isolated subjective cognitive complaints (SCCs) or mild cognitive impairment (MCI; defined as test performance 1.5 SD below age, sex and education-level norms) while not demented (Clinical Dementia Rating [CDR] <1), between January 2011 to June 2014. Participants were followed every 6 months for 5 years[12].

***NFL profiling:*** NFL profiling was carried out using Simoa™ NF-light kit on a Quanterix H1 analyser (Quanterix, MA, USA). Intra CV: 12.7%

***Genotyping and imputation:*** Pre-imputation QC includes removal of SNPs with MAF<0.01, callrate<0.98 and HWE<0.001; Removed samples with callrate<0.05, heterozygosity beyond 3SD, failed sex-check using genotype data of X-chromosome, relates sample based on IBD (pi_hat>0.1875), PCA outliers beyond 6SD of PC1 and PC2. Imputation: HRC.r1.1.2016 (predominantly European Ancestry), phasing using Eagle, Michigan Imputation Server.

**VESTA cohort**

VETSA is a longitudinal behavior genetics study of cognitive and brain aging. The VETSA study design has several key characteristics. First, the sample has a narrow age range (~10 years), allowing for examination of individual differences in aging trajectories. Second, the initial assessment was in midlife (mean age 56; range 51-60), which provides a baseline for the transition to older age. Third, data previously collected on VETSA was obtained; of particular importance is a test of general cognitive ability administered at average age 20 and repeated in each wave of the study. Participants are members of the Vietnam Era Twin Registry, which is housed at the VA Puget Sound Health Care System in Seattle, WA, USA. All of the twins served the US military at some time during the Vietnam era (1965-1975). A 1992 study sought to recruit all Registry twins. It enrolled approximately 8000 individuals, including approximately 3300 twin pairs. VETSA participants were randomly recruited from those 3300 pairs. Eligibility for inclusion was based only on being 51-59 years old at the time of recruitment and willingness of both twins in a pair to participate. Both members of a pair did not need to participate to be included in wave 2 or wave 3. The average interval between assessment was approximately 6 years. Additional participants, including attrition replacement participants, were included at waves 2 and 3. Subsets have multi-modal MRI and neuroendocrine data. Data collection includes questionnaires filled out at home plus a daylong series of assessments. These include cognitive/neuropsychological assessment of multiple cognitive domains, personality and psychosocial assessments, and health/medical assessments. There are approximately 55% MZ and 45% DZ twins in the cohort. For cognitive, psychosocial, and health/medical data, there are 1291 individuals at wave 1, 1207 at wave 2 data, and 1196 at wave 3. Brain MRIs were obtained from 546 individuals at wave 1, 452 at wave 2, and 525 at wave 3. At wave 1 only, salivary cortisol, testosterone, and DHEAS data were collected on 780 participants.

VETSA participants live throughout the US. The sample is primarily Caucasian (European-American): 86% based on self-report. Only those of European-American ancestry based on genotype data were included in GWAS analyses. The average educational attainment is 13.8 years (SD=2.1). At wave 1, 79% were married, and 78% were employed full-time. Nearly 80% report no combat experience. The sample is similar with respect to health and lifestyle characteristics to American men in their age range based on US Center for Disease Control and Prevention data[13-15]. This study was performed under appropriate oversight, including review boards at UC San Diego, Boston University, and VA Puget sound.

**NFL quantification: Nfl:** High throughput bioassays platforms or single analyte assays using the Quanterix Simoa HD-1 were used in this study. These human-specific immunoassays have been documented for measurements of these components in human plasma, as purified in Winston et al., 2016 and many other papers from the laboratory of Dr. Robert Rissman at UC San Diego. These assays are used routinely in the Rissman lab and all assays were performed according the manufacturer instructions.

**Genotyping and imputation:** The methods for genotyping, QC, and imputation have been described in detail elsewhere. See Logue et al. Mol Psych 2019 (PMCID: PMC6110977) for the details. In short, genotypes for VETSA participants were generated using the Illumina HumanOmniExpress-24 v1.0A BeadChip. Imputation was performed with 1000 genomes Phase 3 Reference data using MiniMac on the

Michigan Imputation Server (https://imputationserver.sph.umich.edu). The analysis was done with RMW-RareMetalWorker v 4.13.7. Covariates included age, PCs 1-3, -- RMW already incorporates adjustments which take twinness into account including MZ/DZ relationships.

**The Atherosclerosis Risk in Communities (ARIC) study**

The ARIC study, sponsored by the National Heart, Lung, and Blood Institute (NHLBI), is a prospective, bi-racial population-based study of atherosclerosis and cardiovascular diseases in 15,792 adults (11,478 non-Hispanic whites; 8,710 women) aged between 45 and 64 years at the baseline examination in 1987-1989. Participants were randomly selected and recruited from communities in Suburban Minneapolis, Minnesota; Washington County, Maryland; Forsyth County, North Carolina. In Jackson, Mississippi, only black residents were enrolled. Details about the ARIC study design and examination procedures have been previously published[16]. The ARIC study has been reviewed and approved by the Institutional Review Boards at all participating institutions. All participants provided written informed samples. This study uses data for participants who provided a blood draw at visit 3 (1993-1995) and who completed brain MRI and cognitive testing at visit 5 (2011-2013).

**NFL quantification:** NFL levels were quantified using the the Quanterix 4 plex and assays at University of Vermont by Russ Tracy.

**Genotyping:** Genotyping was performed using the Affymetrix GeneChip SNP Array 6.0 at Broad institute USA. Pre-imputation quality control include sample call rate > 0.95, and retaining SNP with call rate > 0.95, MAF > 0.01 and HWE p-value > $10^{-5}$. Imputation was performed using IMPUTE2 software on 1000G phase1 v3 reference panel.

**CARDIA**

The CARDIA Study is a multisite population-based study conducted across four field centers: Birmingham, Alabama; Chicago, Illinois; Minneapolis, Minnesota; and Oakland, California. The study was established by the National Heart, Lung, and Blood Institute in 1984-1985. Black and White adults, between the ages of 18 to 30 years at the time of enrollment, were recruited. Participants have completed up to 9 examinations over 30 years. At the $25^{th}$ year examination (2010-2011), participants had a blood draw and a subsample completed the brain MRI sub-study.  This study uses data for this subset of participants.

The CARDIA study is a prospective, multi-center investigation of the natural history and etiology of cardiovascular disease in African Americans and whites 18-30 years of age at the time of initial examination. The initial examination included 5,115 participants selectively recruited to represent proportionate racial, gender, age, and education groups from four communities: Birmingham, AL; Chicago, IL; Minneapolis, MN; and Oakland, CA. Participants from the Birmingham, Chicago, and Minneapolis centers were recruited from the total community or from selected census tracts. Participants from the Oakland center were randomly recruited from the Kaiser-Permanente health plan membership. From the time of initiation of the study in 1985-1986, eight follow-up examinations have been conducted at years 2, 5, 7, 10, 15, 20, 25, and 30. Details of the study design have been published elsewhere[17]. The CARDIA study has been reviewed and approved by the Institutional Review Boards at all participating institutions. At the $25^{th}$ year examination (2010-2011), participants had a blood draw and a subsample completed the brain MRI sub-study.  This study uses data for this subset of participants.

**NFL quantification:** NFL levels were quantified using the the Quanterix 4 plex and assays at University of Vermont by Russ Tracy.

**Genotyping:** Genotyping was performed using the Affymetrix 6.0 SNP array at Broad institute USA. Pre-imputation quality control include sample call rate > 0.95, and retaining SNP with call rate > 0.95, MAF > 0.01 and HWE p-value > $10^{-5}$. Imputation was performed using BEAGLE version 3.3.2 software on 1000G phase1 v3 reference panel.

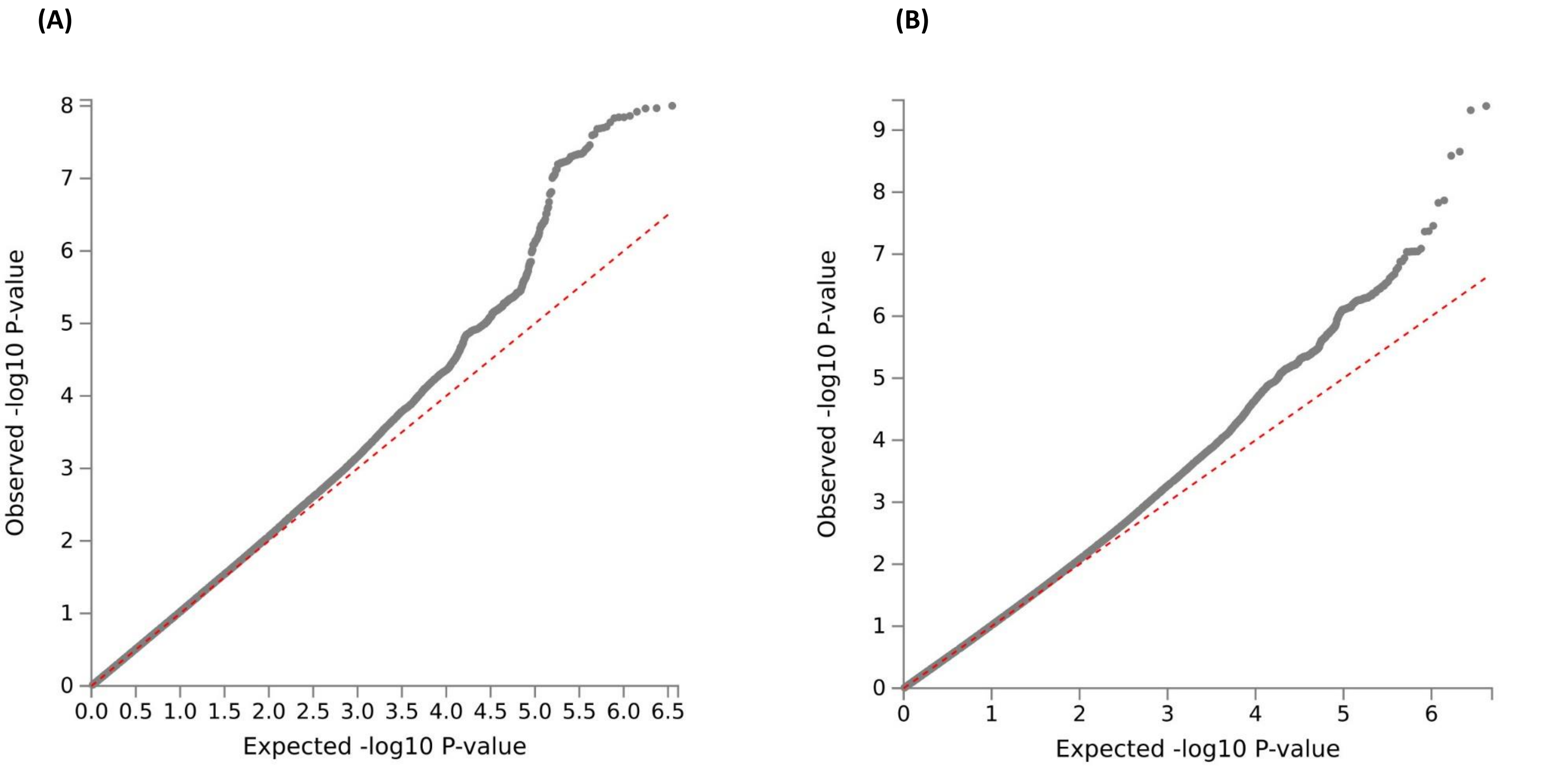


**Supplementary figure 1:** Quantile-Quantile (Q-Q) plot for GWAS meta-analysis for European ancestry (A) and African American Ancestry (B). The grey dotted line represent the distribution of observed –log10(p-values) against the theoretical model distribution of expected log10(p-values). The red dotted line represents the theoretical model distribution of expected –log10(p-values) under the null distribution.

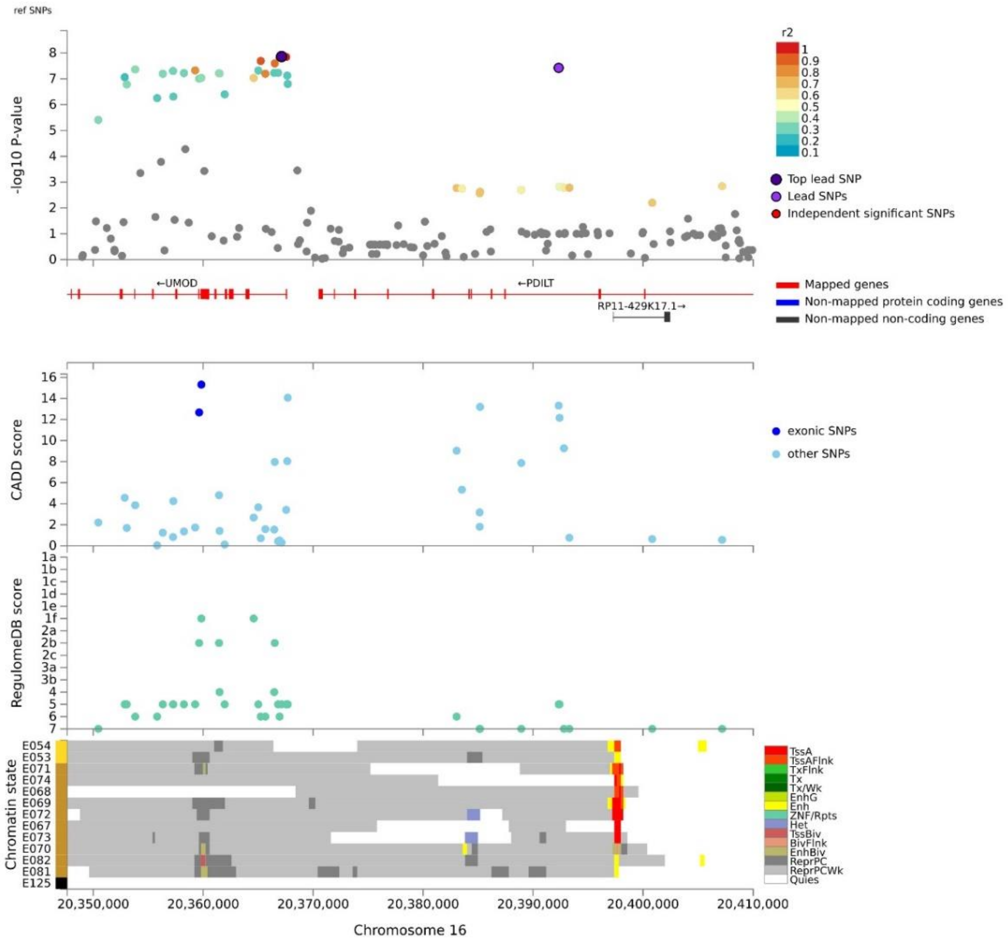


**Supplementary figure 2:** Regional plot showing the top lead genetic variant inside *UMOD* gene, their GWAS *P-value* as –log10, Combined Annotation Dependent Depletion (CADD) score, Regulome Data Base score and expression Quantitative loci (eQTLs). The regional plot with annotation information is generated using the Functional Mapping and Annotation of Genome-Wide Association Studies (FUMA) platform.

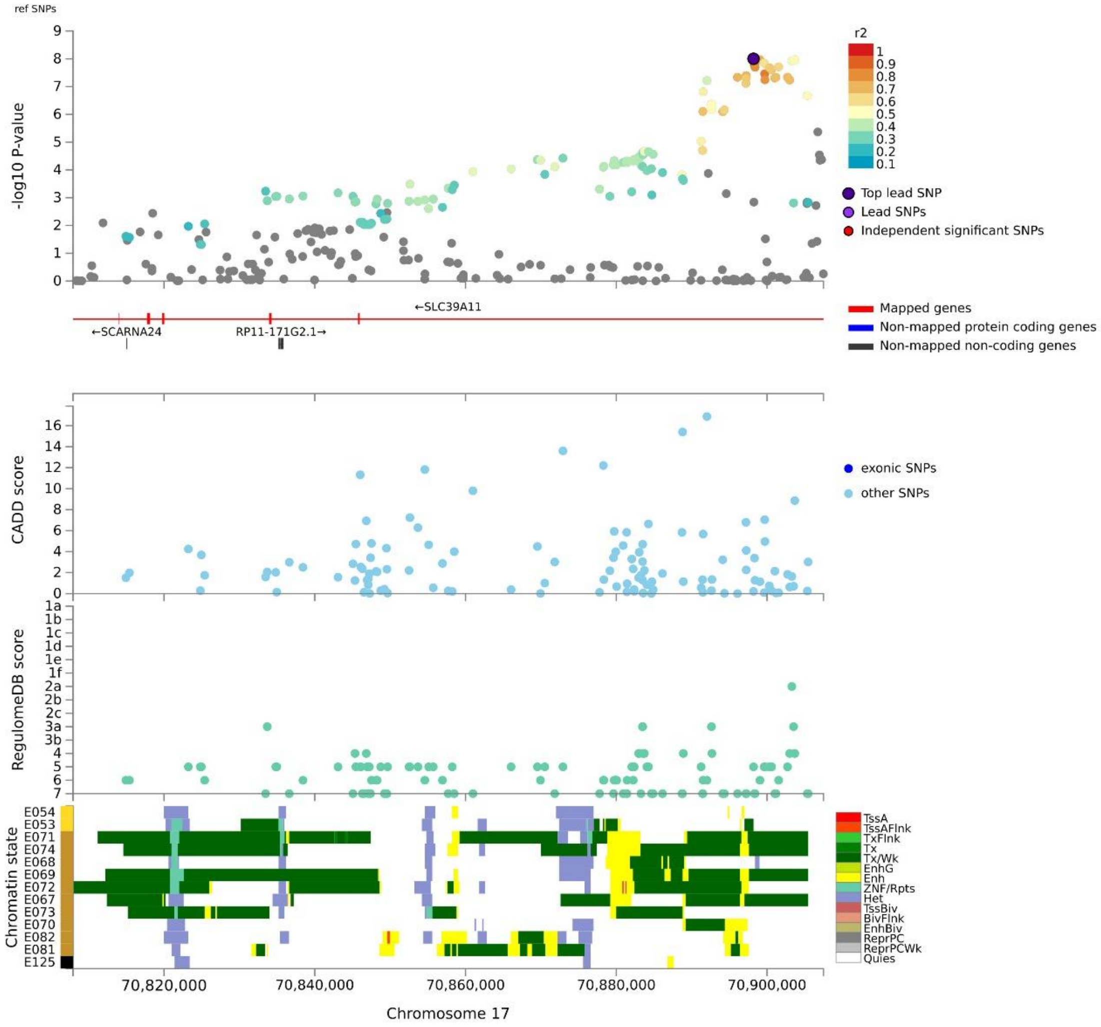


**Supplementary Figure 3:** Regional plot showing the top Lead SNP inside SLC39A11 gene, their GWAS P-value as –log10, Combined Annotation Dependent Depletion (CADD) score, Regulome Data Base score and expression Quantitative loci (eQTLs). The regional plot with annotation information is generated using the Functional Mapping and Annotation of Genome-Wide Association Studies (FUMA) platform.

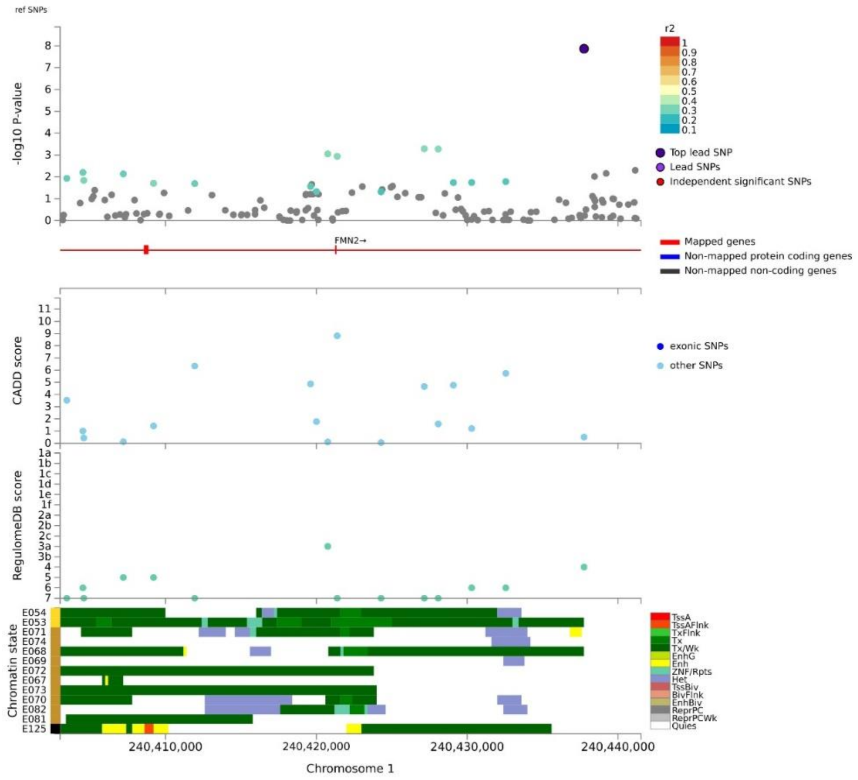

**Supplementary Figure 4:** Regional plot for locus 1q43 in African-American ancestry, their GWAS P-value as –log10, Combined Annotation Dependent Depletion (CADD) score, Regulome Data Base score and expression Quantitative loci (eQTLs). The regional plot with annotation information is generated using the Functional Mapping and Annotation of Genome-Wide Association Studies (FUMA) platform**.**

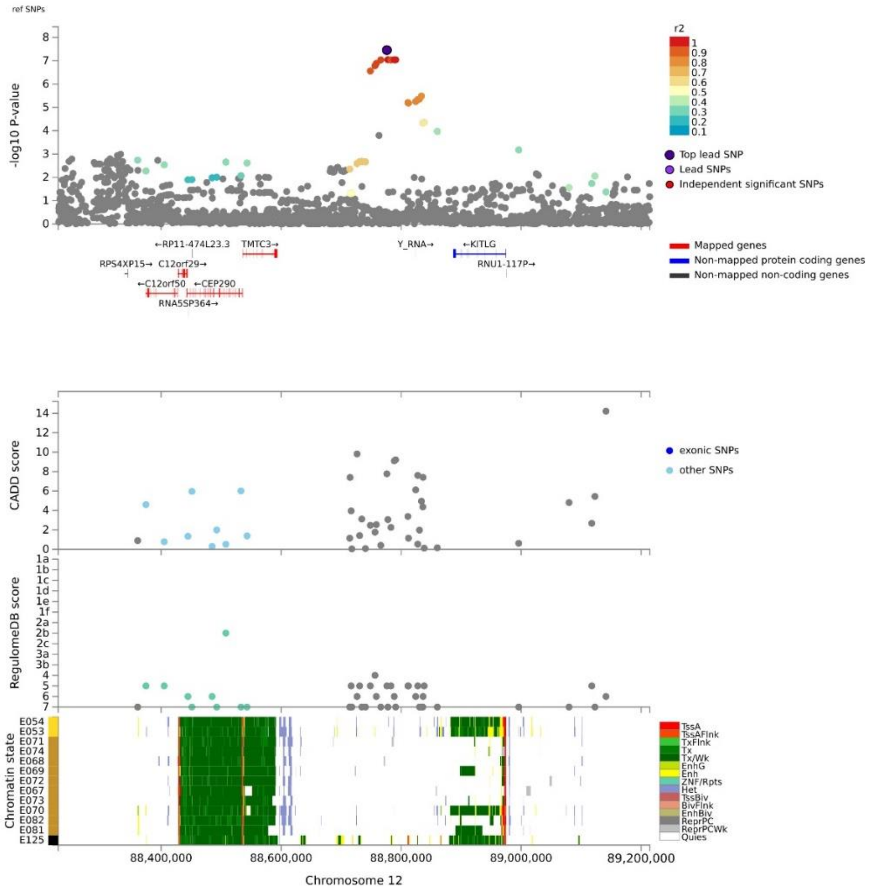


**Supplementary Figure 5:** Regional plot for locus 12q14 in African-American ancestry, their GWAS P-value as –log10, Combined Annotation Dependent Depletion (CADD) score, Regulome Data Base score and expression Quantitative loci (eQTLs). The regional plot with annotation information is generated using the Functional Mapping and Annotation of Genome-Wide Association Studies (FUMA) platform.

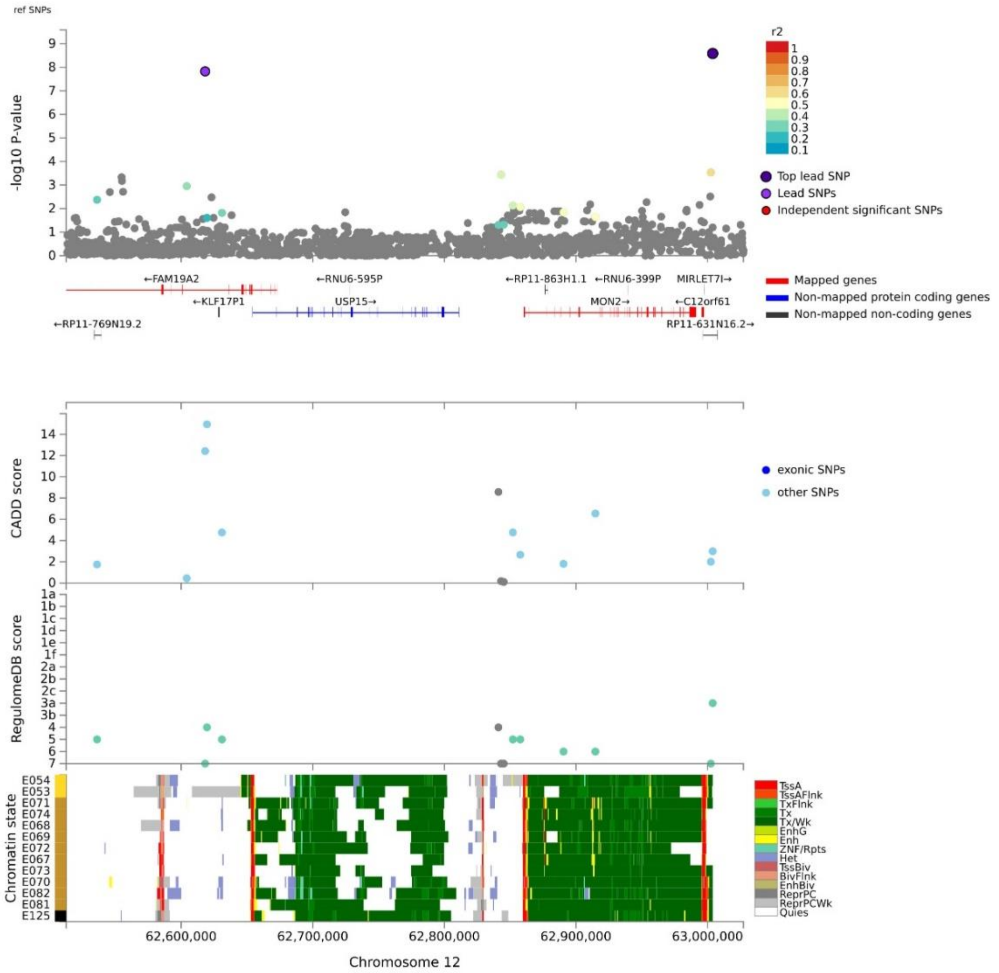


**Supplementary Figure 6**: Regional plot for locus 12q21 in African-American ancestry, their GWAS P-value as –log10, Combined Annotation Dependent Depletion (CADD) score, Regulome Data Base score and expression Quantitative loci (eQTLs). The regional plot with annotation information is generated using the Functional Mapping and Annotation of Genome-Wide Association Studies (FUMA) platform.
